# Human cranial bone-derived mesenchymal stem cells cultured under microgravity can improve cerebral infarction in rats

**DOI:** 10.1101/2023.12.15.23300058

**Authors:** Masashi Kuwabara, Takafumi Mitsuhara, Masataka Teranishi, Takahito Okazaki, Masaaki Takeda, Daizo Ishii, Hiroshi Kondo, Kiyoharu Shimizu, Masahiro Hosogai, Takeshi Hara, Yuyo Maeda, Tomoyuki Kurose, Yumi Kawahara, Louis Yuge, Nobutaka Horie

**Affiliations:** Department of Neurosurgery, Graduate School of Biomedical and Health Sciences, Hiroshima University, Hiroshima, Japan; Division of Bio-Environmental Adaptation Sciences, Graduate School of Biomedical and Health Sciences, Hiroshima University, Hiroshima, Japan; Space Bio-Laboratories Co., Ltd., Hiroshima, Japan

**Keywords:** mesenchymal stem cells, simulated microgravity, cerebral infarction, human cranial bone, transplantation

## Abstract

**Background:** Transplantation of human cranial bone-derived mesenchymal stem cells (hcMSCs) in patients with central nervous system disorders may improve motor function through high neurotrophic factor expression. However, the effects of hcMSCs cultured under microgravity (MG) conditions on cerebral infarction remain unclear. Thus, this study investigated the transplantation effects of hcMSCs cultured in a simulated MG environment on cerebral infarction model rats.

**Methods:** For immunohistological analysis and neurological function evaluation, hcMSCs cultured in normal gravity (1G) or MG environment were transplanted in rats 1 day after inducing cerebral infarction. The expression of endogenous neurotrophic; axonal, neuronal, and synaptogenic; angiogenic; and apoptosis-related factors in infarcted rat brain tissue was examined by real-time polymerase chain reaction (PCR) and western blotting analyses 35 days after stroke induction. MicroRNAs (miRNAs) of hcMSCs cultured under 1G or MG environments were sequenced.

**Results:** Neurological function was significantly improved after transplantation of hcMSCs from the MG group compared with that from the 1G group. Protein expressions of nerve growth factor, fibroblast growth factor 2, and synaptophysin were significantly higher in the MG group than in the 1G group, whereas sortilin 1 expression was significantly lower. MiRNA analysis revealed that genes related to cell proliferation, angiogenesis, neurotrophy, anti-apoptosis, neural and synaptic organization, and cell differentiation inhibition were significantly upregulated in the MG group. In contrast, genes promoting microtubule and extracellular matrix formation and cell adhesion, signaling, and differentiation were downregulated.

**Conclusions:** These results indicate that hcMSCs cultured in an MG environment may be a useful source of stem cells for the recovery of neurological function in cerebral infarction patients.

## INTRODUCTION

Functional disorders of the central nervous system (CNS) such as cerebral infarction are clinically difficult to cure.^1–3^ Given this scenario, cell therapy involving cells such as mesenchymal stem cells (MSCs) has garnered substantial interest as a novel approach for treating CNS diseases. MSCs can have potential clinical applications in cell-based therapies, as they can be isolated from various tissues and autologously transplanted, have self-renewal and multilineage differentiation potential and low immune rejection risk, and pose few ethical issues.^1,4^ MSCs can be obtained from diverse human tissues, including bone marrow, fat, dental pulp, and umbilical cord, and their characteristics may vary depending on the tissue of origin.^5,6^

Our research group has focused on the neural crest-derived cranial bone as a new source of MSCs and has been working with human cranial bone-derived MSCs (hcMSCs). Moreover, we have previously demonstrated that cMSCs have higher neurogenic performance than bone marrow-derived MSCs (bMSCs).^3,7^ Additionally, using a cerebral infarction model, we have shown that compared with transplanted human bMSCs (hbMSCs), transplanted hcMSCs provide neuroprotection by highly expressing neurotrophic factors, thereby significantly improving motor function.^2^

The environment in which MSCs are cultured is one of the most important factors affecting the therapeutic effect of MSC transplantation, especially gravity, the most familiar and important environmental factor for humans.^1,8–10^ In fact, recent studies have reported that culturing cells in different gravity environments can result in different therapeutic efficacies.^10,11^

However, the effects of hcMSCs cultured in simulated microgravity (MG) environments on cerebral infarction, including the mechanisms involved, remain unclear. Thus, in this study, we investigated the effects of hcMSCs cultured in different gravity environments on cerebral infarction rat models using both in vitro and in vivo experiments to determine the differences in their transplantation effects.

## METHODS

### Ethical statements

All experiments involving the care or use of animals were conducted according to the guidelines of the National Institutes of Health, the Animal Testing Committee of Hiroshima University, and the Animal Testing Facility of the Hiroshima University Natural Science Support Center, as well as the ARRIVE guidelines (https://arriveguidelines.org). All research designs were approved by the Animal Experimentation Committee of Hiroshima University (approval number: A19-79). All experimental protocols were approved by the Institutional Review Board of Hiroshima University (approval number: E2010-0379). Written informed consent was obtained from patients for skull sampling in accordance with Hiroshima University Hospital guidelines.

### hbMSC and hcMSC isolation and culture

hbMSCs were purchased from Lonza Japan Ltd. (Tokyo, Japan) and cultured in a normal gravity (1G) environment to serve as control. Human cranial bone samples were obtained from bone fragments, as described previously.^1,2,7^ The bone fragments were formed while removing the sphenoid ridge of the frontotemporal bone during craniotomy surgery and were originally intended to be discarded. Because the skull samples were harvested from free bone fragments obtained during the decompression craniotomy of stroke patients, no additional invasive procedures were required for MSC harvesting.

Human cranial bone samples were seeded in 90-mm tissue culture dishes (Sumitomo Bakelite Co., Tokyo, Japan). The dishes were incubated in a humidified environment at 37°C and 5% CO_2_, and the medium was changed every 3 days. Cell adhesion within the dish was observed. When 80% confluence was reached, several passages were made; adherent cells were collected and used as hcMSCs.

### Cell culture in MG environment

Cell culture in the MG environment followed our previously described method.^1,9,10,12^ Briefly, hcMSCs were seeded (3.5 × 10^3^ cells/cm^2^) in culture flasks (Corning Inc., Corning, NY, USA). After 24 h, cells were divided into two groups: 1G and MG. The MG group was cultured in a device that simulates an MG environment, “Gravite^®^” (Space-Bio-Laboratories Co. Ltd., Hiroshima, Japan), and after reaching 80% confluence, the adherent cells were collected. This device can rotate the cells around two axes to minimize the cumulative gravity vector at the device’s center, creating a 10-3G environment similar to that of the International Space Station (ISS).

### Flow cytometry analysis

The aliquots containing 1 × 10^5^ hcMSCs cultured under 1G and MG were incubated with fluorochrome-labeled [fluorescein isothiocyanate (FITC) or phycoerythrin (PE)] antibodies specific for human CD34 (Santa Cruz Biotechnology, Dallas, TX), and CD44, CD45, CD73, CD90, and CD105 (Biolegend Col., San Diego, CA, USA). FITC mouse IgG1 and PE mouse IgG1 (Biolegend) were used as isotype controls. Data collection and analyses were done using FACSVerse (BD Biosciences, San Jose, CA, USA).

### Micro RNA (miRNA) sequencing analysis

miRNA library preparation, sequencing, mapping, gene expression, and gene ontology (GO) enrichment analysis were performed by DNAFORM (Yokohama, Kanagawa, Japan). Passage 3 hcMSCs cultured in 1G and MG environments were collected and total RNA was extracted according to the manufacturer’s protocol. The quality and quantity of extracted miRNA were assessed using QuantiFluor RNA System (Promega) and Agilent Small RNA kit of BioAnalyzer 2100 System (Agilent Technologies), according to the manufacturers’ instruction. miRNA library was prepared using QIAseq miRNA Library Kit (Qiagen) following the manufacturer’s instructions. Differential gene expression analysis and GO analysis were performed using DESeq2 (ver. 1.18.1) and clusterProfiler (ver. 3.6.0), respectively.

### Induction of cerebral infarction in rats

Considering the minimum number of samples required for the study, Middle cerebral artery occlusion (MCAO) was performed on 48 adult, female Sprague–Dawley (SD) rats (body weight, 250–300 g). After anesthesizing the rats by isoflurane inhalation, a 4-0 nylon thread with a silicone-coated tip (403745PK10; Doccol Co., Sharon, MA, USA) was inserted into the internal carotid artery and the middle cerebral artery was occluded at its origin, based on previous methods.^2,3^ After 90 min of occlusion, the nylon threads were removed and reperfusion was performed. The infarcted areas were confirmed by magnetic resonance imaging (MRI) and hematoxylin-eosin (HE) staining 1 and 35 days after MCAO, respectively (Figure 1).

**Figure 1.**
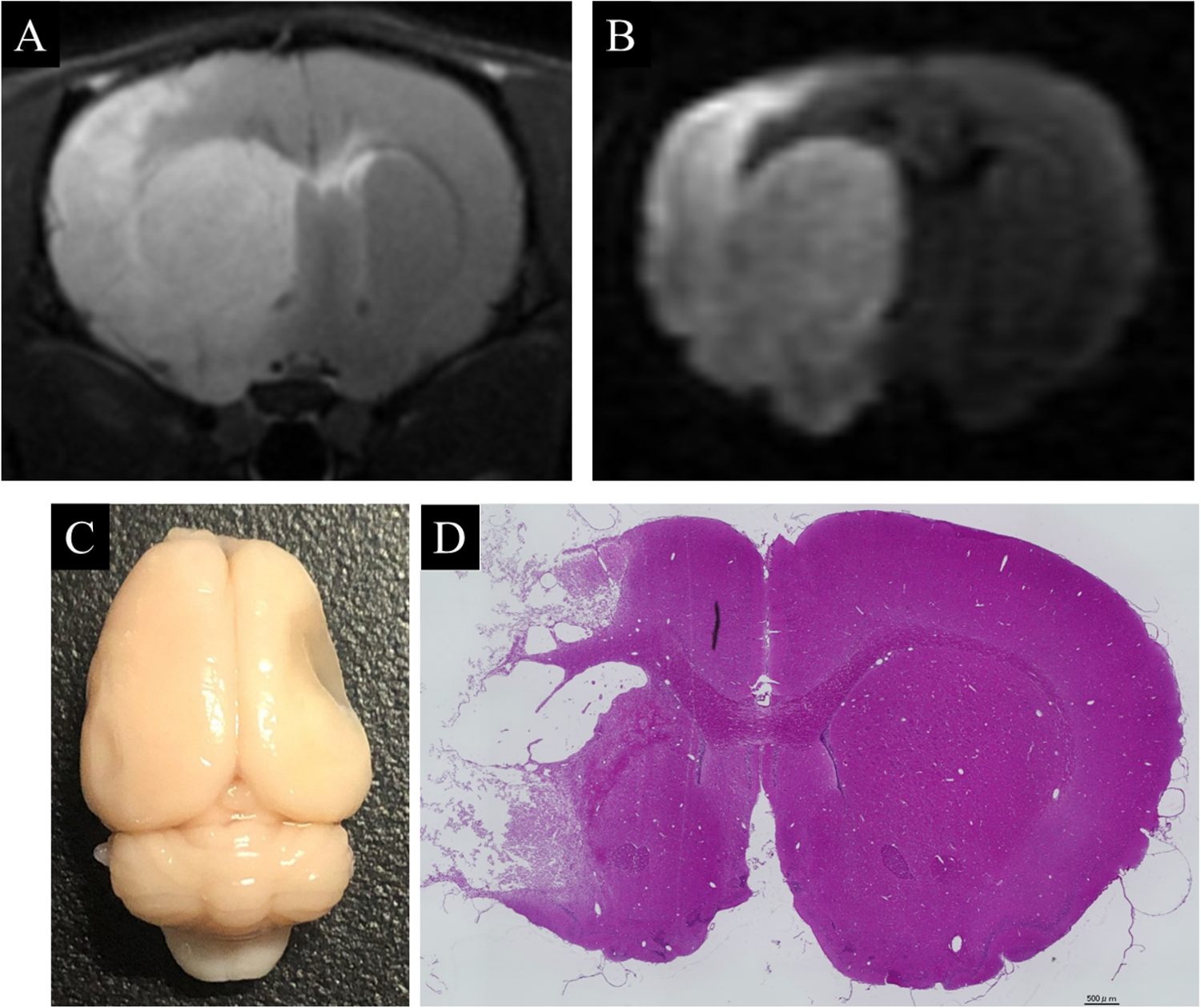
The area of cerebral infarction was confirmed by various methods: T2-weighted image (A) and diffusion-weighted image (B) of magnetic resonance imaging (MRI) on day 1 after middle cerebral artery occlusion (MCAO) showed that the cerebral infarction was visualized as a high-intensity area. On day 35, the area of cerebral infarction was confirmed by macroscopic brain (C) and hematoxylin-eosin (HE) staining (D). Scale bars, 500 μm.

### MRI evaluation

Rats were anesthetized by isoflurane inhalation and scanned in the supine position a day after the MCAO procedure. Respiratory movements of the chest wall in rats in the imaging coil were monitored. The imaging sequence consisted of diffusion-weighted image (DWI) and T2-weighted image (T2WI) evaluated in coronal slices at the level of the caudate–putamen complex. The area showing high-intensity signal in DWI and T2WI was diagnosed as the cerebral infarction area.

### Cell transplantation

After MCAO, rats were divided into the following three groups: group injected with only phosphate-buffered saline as control (PBS group, n = 9), group with transplanted hcMSCs cultured under 1G environment (1G group, n = 9), and group with transplanted hcMSCs cultured under MG environment (MG group, n = 10). Passage 3 hcMSCs (1 × 10^6^ cells/300 μL of PBS) cultured in 1G and MG environments were used for transplantation and were injected in rats via the tail vein in the 1G and MG groups, respectively, 1 day after MCAO. All rats were immunosuppressed by intraperitoneal administration of cyclosporine A (10 mg/kg) daily starting from the day before MCAO.^2,13^

### Assessment of neurological function

The neurological function of the cerebral infarction model rats was evaluated using the modified neurological severity score (mNSS). The mNSS is a scale of motor, sensory, balance, and reflex tests ranging from 0 to 18, with higher scores indicating more severe neurological symptoms.^14^ To reduce the variability of neurological symptoms in rats in this study, we used rats with mNSS scores of 10–13 points 1 day after MCAO. mNSS was evaluated before MCAO and 1, 3, 7, 14, 21, 28, and 35 days after MCAO in the PBS, 1G, and MG groups. To minimize potential bias, blinded assesments were independently conducted by two evaluators.

### Immunohistological evaluation

After 35 days of cell transplantation, perfusion-fixed brain tissues of rats in the PBS, 1G, and MG groups were embedded in paraffin and sliced into 3–5 µm thick sections. Brain sections were then subjected to primary antibody reactions overnight : rabbit anti-synaptophysin antibody (ab32127, 1:500; Abcam, Cambridge, UK) and mouse anti-Tuj1 antibody (GTX631836, 1:250; Gene Tex), followed by incubation with diluted secondary antibodies: FITC-conjugated goat anti-rabbit IgG antibody (1:100; Sigma-Aldrich) and Rhodamine Red-X-AffiniPure Fab Fragment donkey anti-mouse IgG antibody (1:100; Jackson ImmunoResearch). Stained brain sections were sealed in VECTASHIELD Mounting Medium with DAPI (Vector Laboratories) and observed under a fluorescence microscope (FluoView FV-1000, OLYMPUS).

### Real-time polymerase chain reaction (RT-PCR)

Rats in the PBS, 1G, and MG groups were euthanized under deep anesthesia 35 days after cell transplantation (n = 6). Total RNA from the right cerebral hemisphere, the infarct side, was reverse transcribed using an Isogen RNA Extraction Kit (Nippon Gene, Tokyo, Japan), following the manufacturer’s protocol. RT-PCR was performed with the generated cDNA as a template using the 7900HT Real-Time PCR System (Applied Biosystems, Carlsbad, CA, USA). RT-PCR was performed using TaqMan assays for brain-derived neurotrophic factor (Bdnf), neurotrophic tyrosine kinase receptor type 2 (Ntrk2), fibroblast growth factor 2 (Fgf2), nerve growth factor (Ngf), growth-associated protein 43 (Gap43), synaptophysin (Syp), vascular endothelial growth factor (VEGF), and sortilin 1 (Sort1). β-Actin (Act-b) was used as an internal control. The assay IDs of the TaqMan assay primers used in this study are shown in Table S2.

### Western blotting analysis of cerebral infarct samples

After 35 days of cell transplantation, cerebral infarction samples from rats in the PBS, 1G, and MG groups were collected and homogenized, and total protein was extracted using RIPA lysis buffer (ATTO, Tokyo, Japan) (n = 4). Equal amounts of protein were separated electrophoretically using Mini-PROTEAN® TGX™ precast gels (Bio-Rad, Hercules, CA, USA) and transferred to nitrocellulose membranes (Hybond TM-ECL; GE Healthcare, Little Chalfont, UK). Subsequently, the membranes were incubated with primary antibodies against Bax (ab32503, 1:1000; Abcam), Bcl-XL (ab32370, 1:1000; Abcam, SYP (ab32127, 1:5000, Abcam), and Act-b (ab6276, 1:10,000; Abcam) at 4°C overnight. A secondary antibody reaction was performed using horseradish peroxidase-conjugated anti-rabbit IgG (1:2000, Vector Laboratories) for 60 min at room temperature (25°C ± 5°C). Images were detected using a Versa Doc imaging system (Bio-Rad). Expression levels of target proteins were quantified by measuring the concentration of bands using ImageJ (National Institutes of Health, Bethesda, MD, USA). Act-b was loaded as a control and data were normalized.

### Statistical analysis

Statistical analysis was performed using R (ver. 3.1.2, http://www.r-project.org). One-way analysis of variance with the Bonferroni test was used to compare motor function assessment among the three groups. Gene and protein expression data were evaluated using paired t-tests. A p-value < 0.05 was considered statistically significant.

## RESULTS

### Confirmation of hcMSCs cultured in 1G and MG environments

hcMSCs cultured in 1G and MG environments are shown in Figure 2A-C. Both groups of cells showed spindle and oval shapes without any morphological differences.

**Figure 2.**
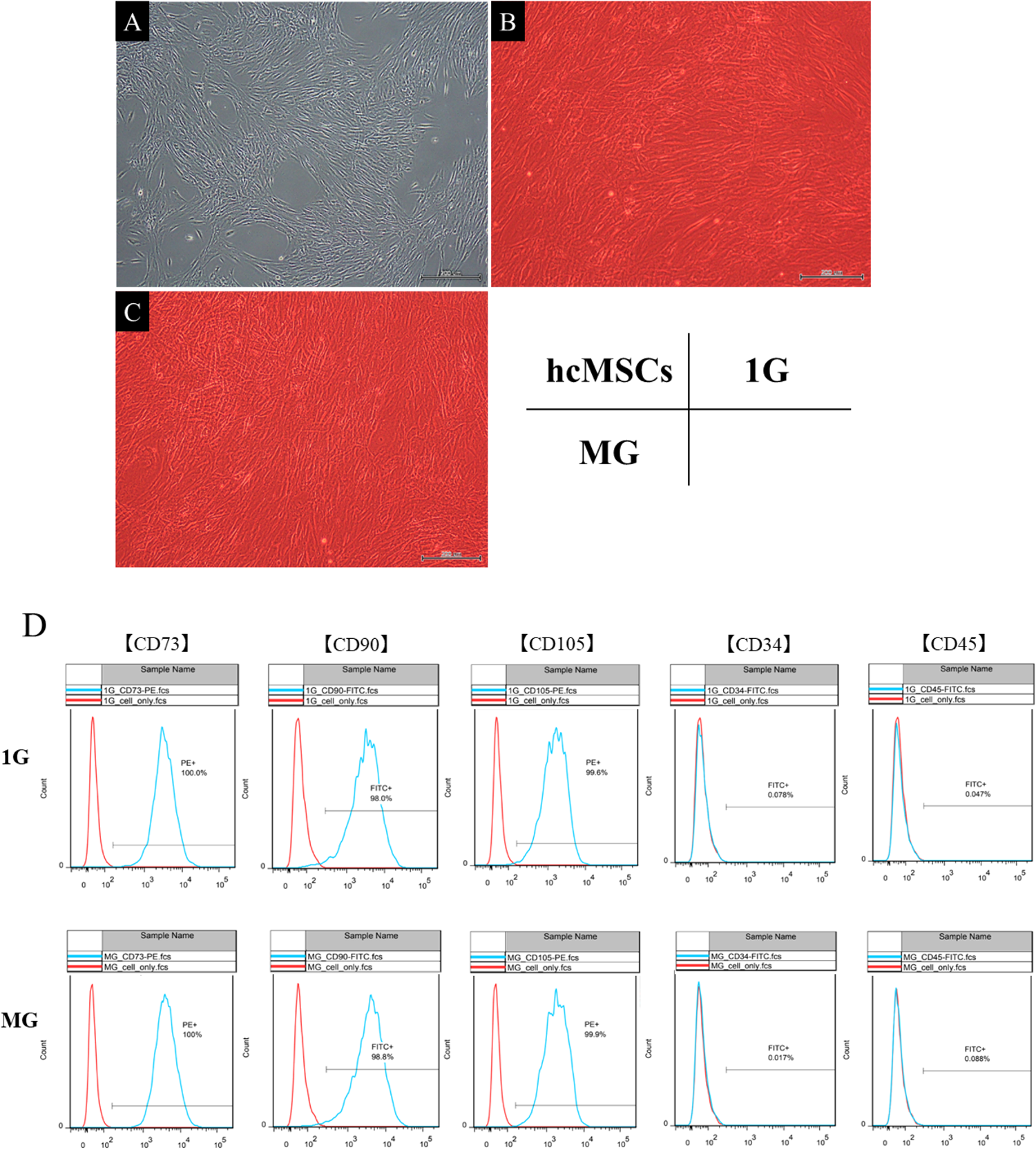
Human cranial bone-derived mesenchymal stem cells (hcMSCs) could be identified as spindle-shaped and oval cells (A). hcMSCs cultured in normal gravity (1G) (B) and microgravity (MG) (C) environments showed no obvious morphological differences. Images in (B) and (C) were captured with the medium in special flasks, so the overall color is red. Flow cytometry analyses showed that hcMSCs cultured under 1G and MG were positive for MSC positive markers (CD73, CD90, and CD105) and negative for MSC negative markers (CD34 and CD45) (D).

### Expression of cell surface markers in hcMSCs cultured in 1G and MG environments

Flow cytometry analyses showed that hcMSCs cultured under both 1G and MG were positive for MSC-positive markers (CD73, CD90, and CD105) and negative for MSC-negative markers (CD34 and CD45) (Table S1, Figure 2D).

### miRNA sequencing analysis

Comparing the miRNAs between hcMSCs cultured in the 1G environment (miRNA-1G group) and those cultured in the MG environment (miRNA-MG group), we found that 324 genes showed significant differences in expression (p < 0.05). Figure 3A shows the GO enrichment analysis results for these genes. Compared with the miRNA-1G group, the miRNA-MG group exhibited significant upregulation of genes related to cell proliferation, angiogenesis, neurotrophy, anti-apoptosis, neural and synaptic organization, and inhibition of cell differentiation. In particular, the fibroblast growth factor 7 (*FGF-7*), epithelial mitogen (*EPGN*), vascular endothelial growth factor-A (*VEGF-A*), angiopoietin 1 (*ANGPT1*), and glial cell-derived neurotrophic factor (*GDNF*) genes were significantly more expressed in the miRNA-MG group than in the miRNA-1G group (p < 0.05). In contrast, genes promoting microtubule and extracellular matrix formation, cell adhesion, and cell differentiation were significantly more downregulated in the miRNA-MG group than in the miRNA-1G group. Specifically, the cathepsin S (*CTSS*), laminin subunit alpha 1 (*LAMA1*), fibrillin 2 (*FBN2*), dynactin subunit 1 (*DCTN1*), and integrin subunit beta 3 (*ITGB3*) genes were significantly less expressed in the miRNA-MG group than the miRNA-1G group (p < 0.05).

**Figure 3.**
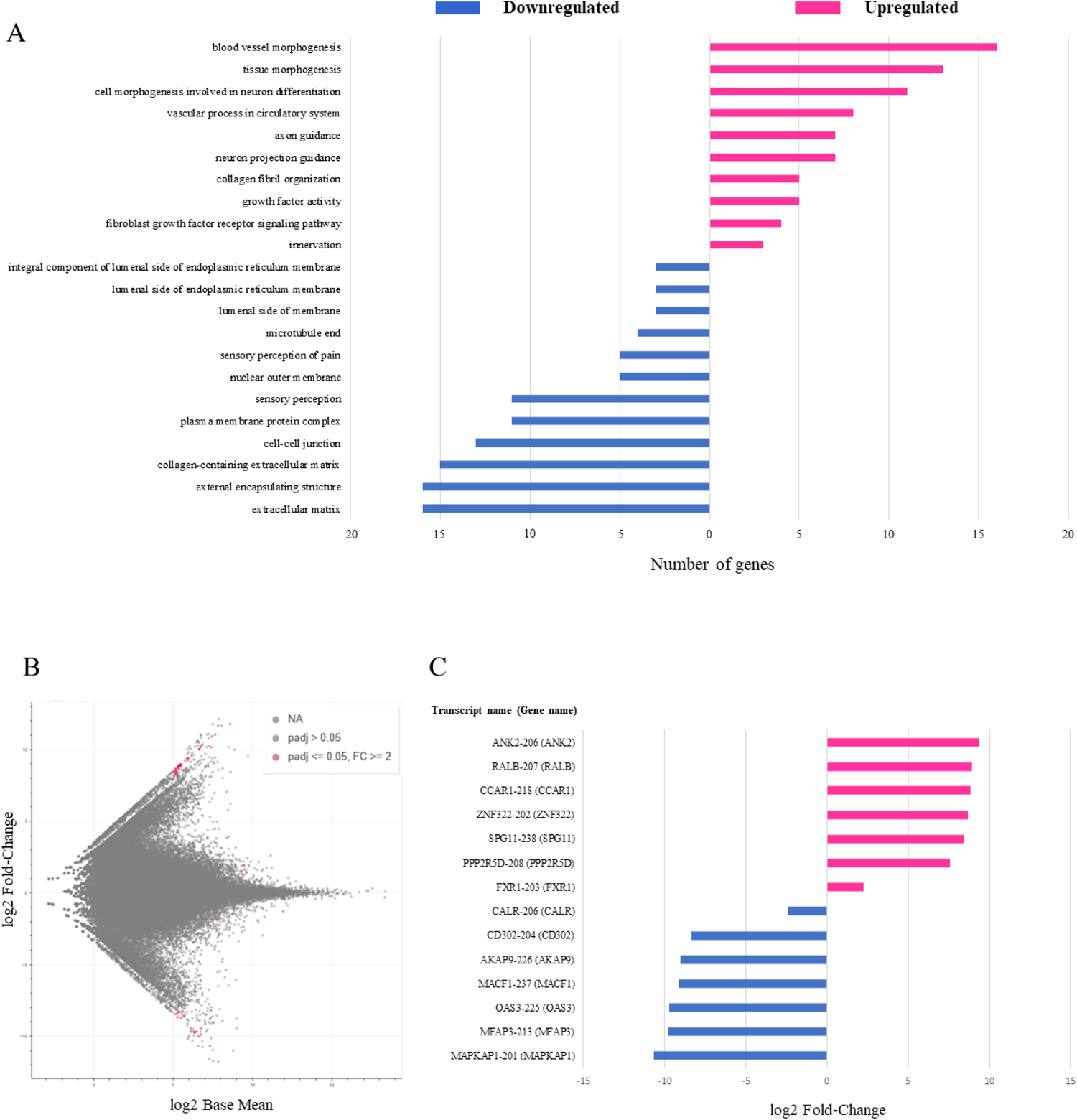
(A) Gene ontology enrichment analysis revealed that genes related to cell proliferation, angiogenesis, neurotrophy, anti-apoptosis, neural and synaptic organization, and cell differentiation inhibition were significantly upregulated in hcMSCs cultured in MG (miRNA-MG group) compared with hcMSCs cultured in 1G (miRNA-1G group). In contrast, genes promoting microtubule and extracellular matrix formation, cell adhesion, and cell differentiation were significantly downregulated in the miRNA-MG group compared with the miRNA-1G group. (B) MA plot showing the comparison between the miRNA transcripts in the miRNA-1G group and those in the miRNA-MG group; 74 transcripts showed significant differences in expression (p-adjust < 0.05). (C) Genes corresponding to each transcript were characterized. Genes inducing cell proliferation, angiogenesis, and anti-apoptosis were significantly more expressed in the miRNA-MG group than in the miRNA-1G group (p-adjust < 0.05). In contrast, genes that promote microtubule and cytoskeleton formation and stabilization, cell adhesion, and cell differentiation were significantly less expressed in the miRNA-MG group than in the miRNA-1G group (p-adjust < 0.05).

Furthermore, comparing the transcripts between the miRNA-1G and miRNA-MG groups, we found that 74 transcripts showed significant differences in expression (p-adjust < 0.05, Figure 3B). The characterization of genes corresponding to each transcript showed that genes inducing cell proliferation and migration, neurite outgrowth, angiogenesis, anti-inflammation, and anti-apoptosis were significantly more highly expressed in the miRNA-MG group than in the miRNA-1G group. In contrast, genes that promote microtubule and cytoskeleton formation and stabilization, cell adhesion, cell signaling, apoptosis induction, and cell differentiation were significantly less expressed (p-adjust < 0.05, Figure 3C) in the miRNA-MG group than in the miRNA-1G group. These results were considerably similar to the results of GO enrichment analysis on genes.

### Evaluation of neurological function in cerebral infarction model rats

The mNSS was used to evaluate the neurological function in the PBS, 1G, and MG groups (Figure 4A). Rats with MCAO in the MG group showed significantly improved neurological function compared with those in the PBS group starting from day 3 after transplantation till days 7, 14, 21, 28, and 35. Furthermore, compared with the 1G group, the MG group showed significantly improved neurological function from day 7 after transplantation, with significantly higher neurological recovery on days 14, 21, 28, and 35.

**Figure 4.**
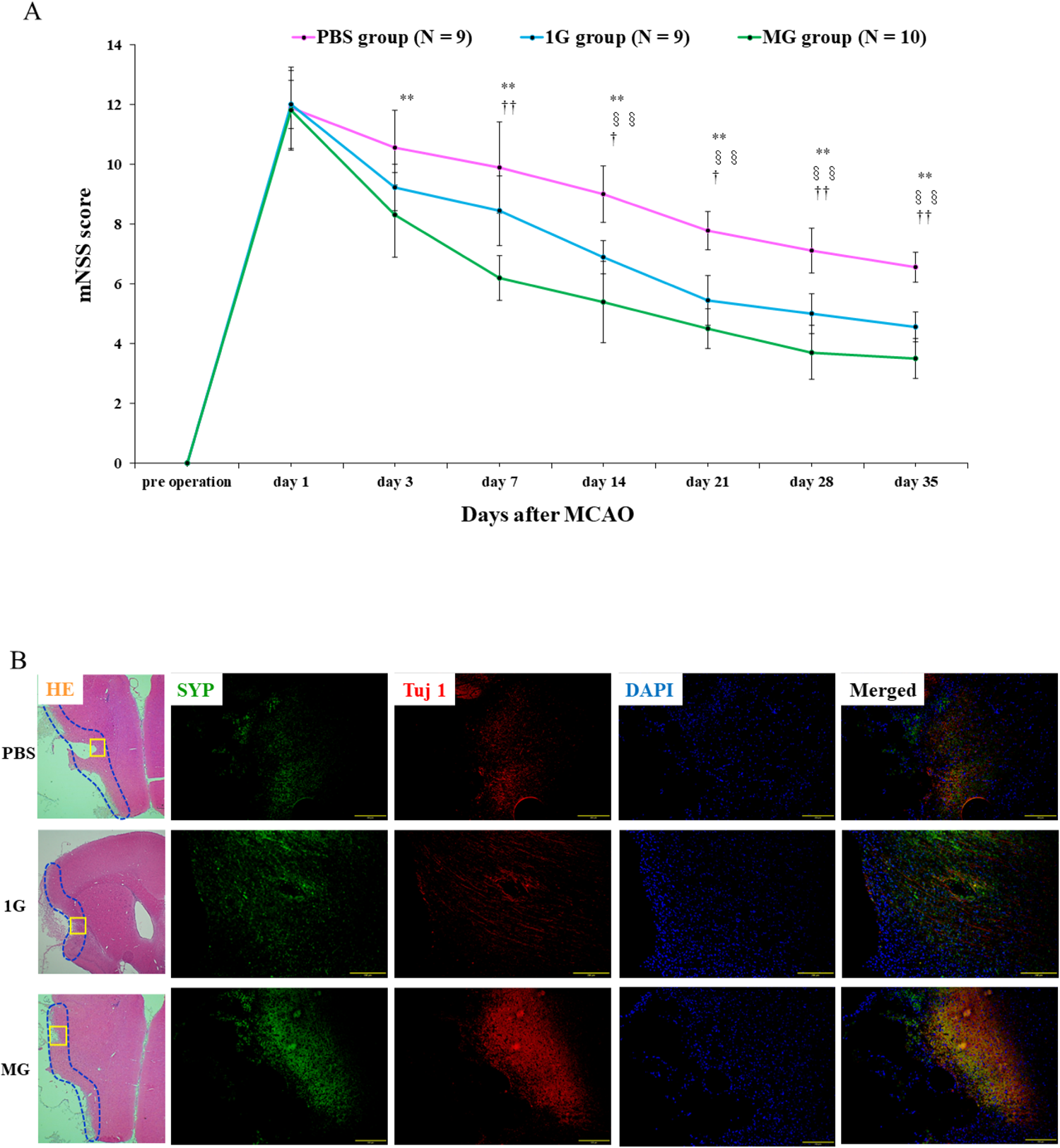
(A) Evaluation of neurological function in cerebral infarction model rats implanted with human cranial bone-derived mesenchymal stem cells (hcMSCs) cultured in normal gravity (1G) and microgravity (MG) environments. The group implanted with phosphate-buffered saline (PBS) served as the control. The MG group had significantly improved neurological function compared with the PBS and 1G groups. Data are presented as the mean ± standard deviation. PBS group, n = 9; 1G group, n = 9; MG group, n = 10. *PBS group versus MG group; †1G group versus MG group; §PBS group versus 1G group. **p < 0.01, †p < 0.05, ††p < 0.01, and §§p < 0.01. (B) Micrographs showing immunofluorescence in the cerebral infarct area in the PBS group (top row), 1G group (middle row), and MG group (bottom row) 35 days after cell transplantation to cerebral infarction model rats. The left column is HE: hematoxylin-eosin staining (orange) and SYP: synaptophysin (green). The middle column is Tuj1 (red). The right column is DAPI (blue) and merged image (black). Scale bars, 200 µm (HE staining: 100 µm). In each group, SYP and Tuj1 were distributed in the ischemic border zone (IBZ), which is the border area of cerebral infarction. In the PBS group, both Tuj1 and SYP were sparsely distributed, whereas the 1G and MG groups transplanted with hcMSCs showed relatively high amounts of Tuj1 and SYP in the IBZ. Furthermore, especially in the MG group, Tuj1 and SYP showed strong staining intensity along the IBZ.

### Immunohistological evaluation

Immunohistological evaluation of the cerebral infarction area in rats in the PBS, 1G, and MG groups 35 days after cell transplantation showed that SYP and a marker for neurons, Tuj1, were distributed in the ischemic border zone (IBZ) of the infarct in each group (Figure 4B). In the PBS group, both Tuj1 and SYP were sparsely distributed, whereas the 1G and MG groups transplanted with hcMSCs showed relatively high amounts of Tuj1 and SYP in the IBZ. Furthermore, in the MG group, Tuj1 and SYP showed strong staining intensity along the IBZ.

### Effects of transplantation of MG environment-cultured hcMSCs on mRNA expression at the cerebral infarction site

The mRNA expression levels in the infarcted brain tissues of the three rat groups were evaluated 35 days after cell transplantation using RT-PCR (Figure 5). The expression levels of Ngf and Fgf2 were significantly higher in the MG group compared with the PBS group. Additionally, the Syp expression level was significantly higher in the MG group than in the PBS and 1G groups. However, the expression of Sort1 was significantly lower in the MG group than in the PBS group. BDNF, Ntrk2, Gap43, and VEGF expressions were not significantly different among the three groups.

**Figure 5.**
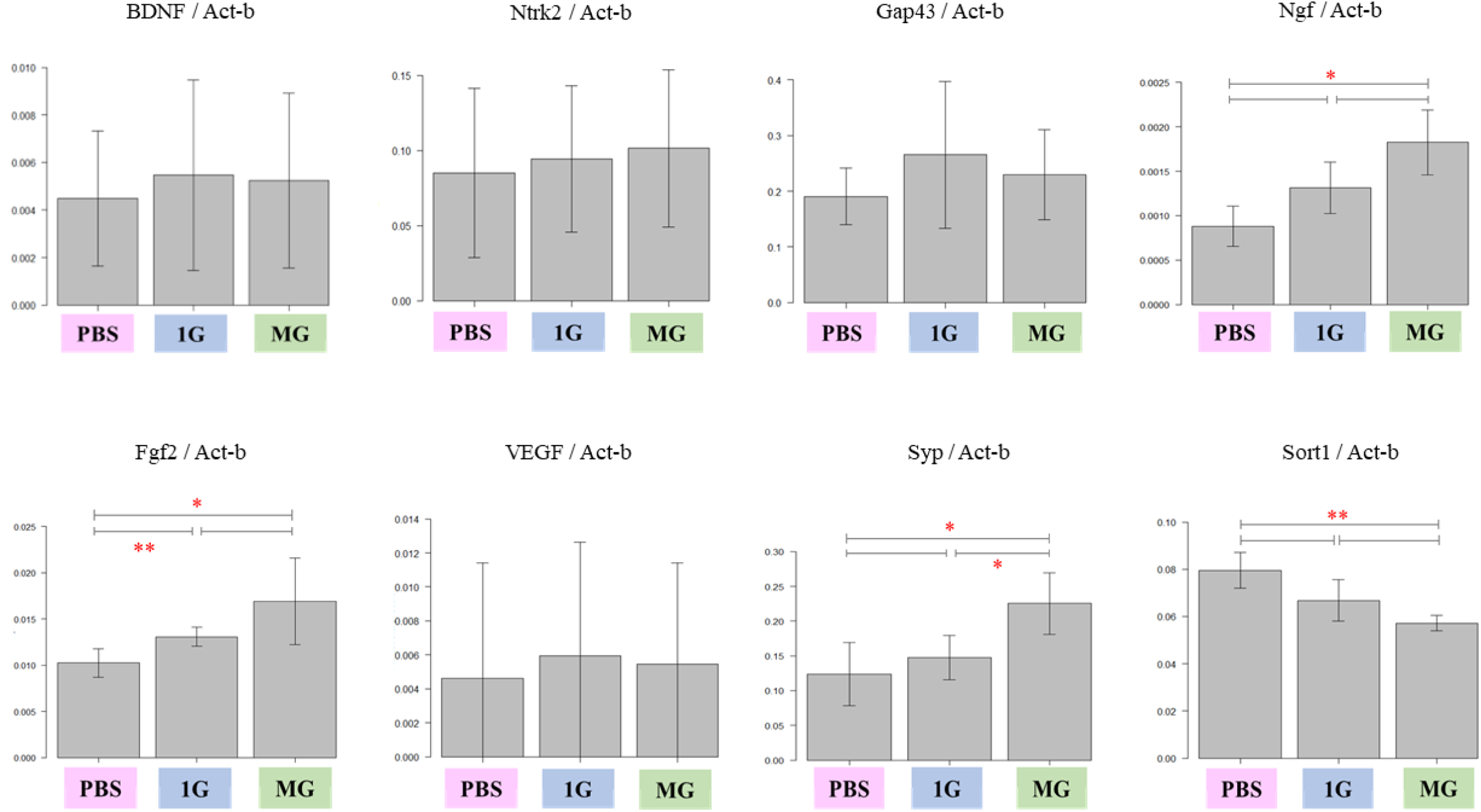
mRNA expression levels in infarcted brain tissue 35 days after 1G- and MG-cultured hcMSC transplantation. Data are represented as bar graphs normalized by β-Actin (Act-b). BDNF: brain-derived neurotrophic factor, Ntrk2: neurotrophic tyrosine kinase receptor type 2, Gap43: growth-associated protein 43, Ngf: nerve growth factor, Fgf2: fibroblast growth factor 2, VEGF: vascular endothelial growth factor, Syp: synaptophysin, Sort1: sortilin 1. n = 6, *p < 0.05, **p < 0.01. The expression levels of Ngf and Fgf2 were significantly higher in the MG group than in the PBS group. Syp expression was significantly higher in the MG group than in the PBS and 1G groups. However, Sort1 expression was significantly lower in the MG group than in the PBS group. Furthermore, BDNF, Ntrk2, Gap43, and VEGF expressions were not significantly different among the groups.

### Effects of transplantation of MG environment-cultured hcMSCs on protein expression at the cerebral infarction site

Protein expression levels in infarcted brain tissue 35 days after transplantation of hcMSCs cultured in the 1G and MG environments were analyzed using western blotting (Figure 6A). Syp expression was significantly higher in MG and 1G rats than in PBS rats (Figure 6D). However, Bcl-2-associated X protein (Bax) or B-cell lymphoma-extra large (Bcl-xL) expression was not significantly different among the three groups (Figure 6B, C).

**Figure 6.**
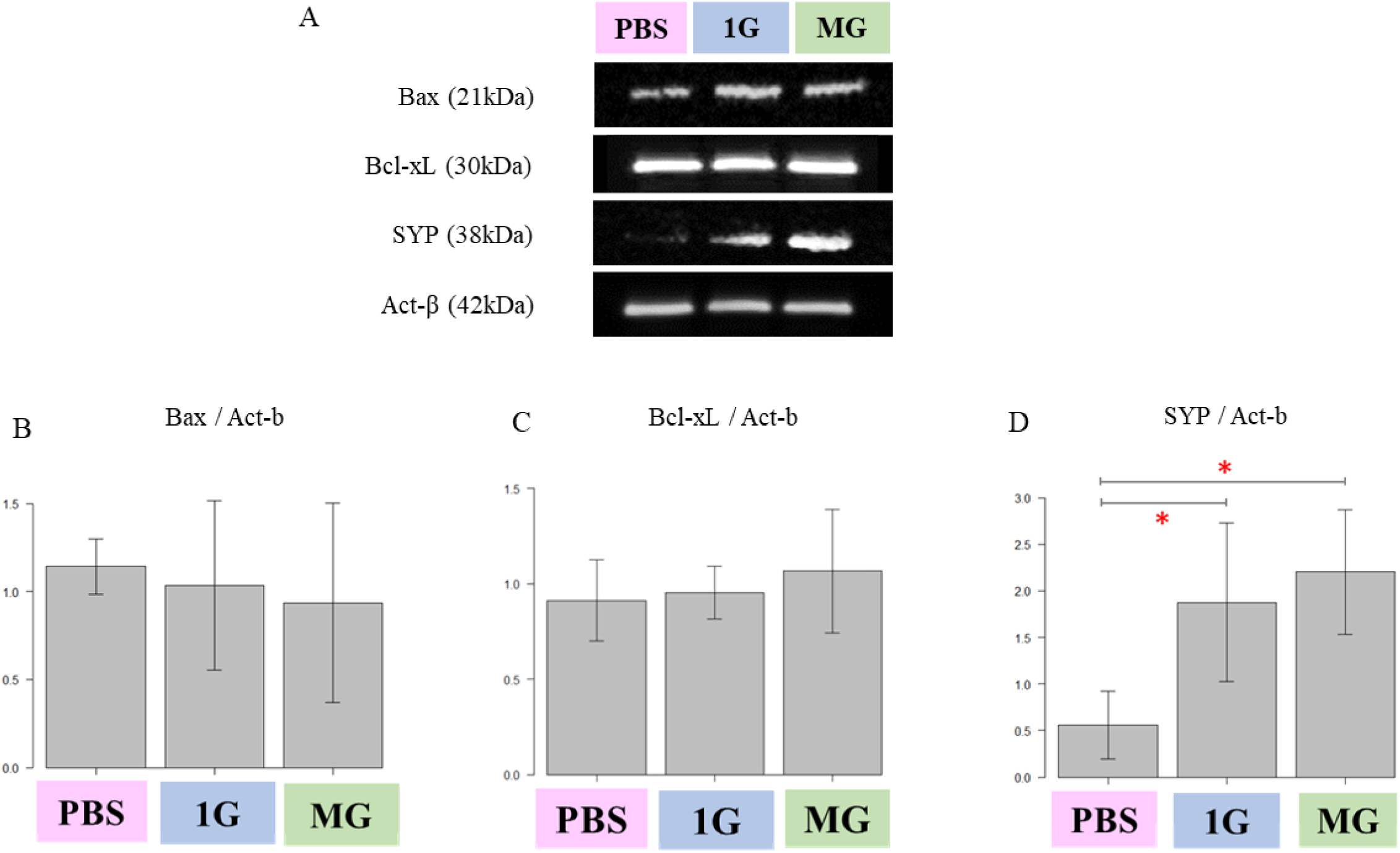
Electrophoretic band images of proteins (A) in infarcted brain tissue 35 days after 1G- and MG-cultured hcMSC transplantation. Bar graphs showing protein levels of Bcl-2-associated X protein (Bax) (B), B-cell lymphoma-extra large (Bcl-xL) (C), and synaptophysin (Syp) (D), normalized by β-Actin (Act-b). n = 4, *p < 0.05. Syp expression was significantly higher in the MG and 1G groups than in the PBS group. However, no significant differences in Bax and Bcl-xL expression were observed among the three groups.

## DISCUSSION

In this study, we confirmed that compared with transplantation of hcMSCs cultured under 1G environment, transvenous transplantation of hcMSCs cultured under MG environment significantly enhanced neurological functional recovery in cerebral infarction model rats. However, studies on the effects of transplanting cells cultured in an MG environment in disease models are limited; to our knowledge, this is the first study to examine such effects of hcMSCs cultured in an MG environment on cerebral infarction.^15^

Previously, our research group successfully established neural crest cell-derived hcMSCs from human skulls and found that hcMSCs have higher transplantation efficacy than hbMSCs.^2,7^ Herein, we expanded on this observation and focused on the environmental conditions under which hcMSCs are cultured. We found that the mRNA expression levels of Ngf and Fgf2 were significantly higher in the MG group than in the PBS group, and Syp expression was significantly higher in the MG group than in the 1G group. On the other hand, Sort1 expression was significantly lower in the MG group than in the PBS group. Similarly, in terms of protein expression levels, Syp expression was significantly higher in the MG and 1G groups than in the PBS group, further supporting the results of RT-PCR. Furthermore, immunohistological examination confirmed the distribution of SYP and Tuj1 in the IBZ of cerebral infarcts, particularly in the MG group, in which Tuj1 and SYP were strongly stained along the IBZ, supporting the results of gene-protein analyses.

The *Fgf2* gene plays an important role in the regulation of cell division, differentiation, and migration and promotes cell proliferation and angiogenesis.^16,17^ Ngf stimulates nerve growth and is associated with the regulation of neurite outgrowth, differentiation, and survival; axon elongation; healing after cell injury; and the development and maintenance of sympathetic and sensory nervous systems.^18,19^ Syp proteins encoded by the *Syp* gene are located in presynaptic vesicles and are important regulators of synapse formation and plasticity; fluctuations in Syp expression during motor neuron injury are an important indicator reflecting motor function loss and recovery.^20,21^ Sort1, on the other hand, promotes neuronal apoptosis and regulates neuronal development.^22^ To summarize, the transplantation of hcMSCs cultured in MG environment significantly altered the expression levels of Fgf2, Ngf, Syp, and Sort1 in the infarcted brain tissues of rats, which had a multifaceted effect on cell proliferation, angiogenesis, axon elongation, wound healing, synaptogenesis, and anti-apoptosis, inducing neural plasticity and significantly influencing neurological function recovery.

The present study focused on the effect of gravity on hcMSCs. In recent years, many experiments have been conducted in space, suggesting that an MG environment is a useful tool for cell culture in cell therapy.^9,10,23,24^ However, since experiments in space are highly expensive, we used “Gravite,” a multi-directional gravity control device developed by our group, to simulate MG.^1,11,25^ In vitro experiments showed that genes related to cell proliferation, angiogenesis, neurotrophy, anti-apoptosis, nerve and synapse organization, and cell differentiation inhibition were significantly upregulated in the miRNA-MG group than in the miRNA-1G group. In contrast, genes promoting microtubule and extracellular matrix formation, cell adhesion, cell signaling, and cell differentiation were significantly downregulated in the miRNA-MG group compared with the miRNA-1G group. Similar results were obtained when transcripts between the miRNA-1G and miRNA-MG groups were compared.

Although the detailed specific mechanisms underlying the various effects of an MG environment on cells, including MSCs, are still unknown, several studies have reported these effects. Experiments examining changes in mouse brain tissue in actual cosmic space have reported increased dendritic spines of motoneurons, increased number and size of axonal mitochondria, changes in synaptic morphology, and increased synaptic transmission.^23,24^ Experiments analyzing changes in bMSCs exposed to an MG environment by spaceflight showed downregulation of cytoskeletal genes and cytoskeleton changes, suggesting that the expression of genes involved in neurogenesis is altered, promoting the activation of neurogenic processes such as nervous system development, differentiating neuron morphogenesis, and increased synaptic transmission.^9,26^ Regarding the effect of MG environment on the cytoskeleton, Ulbrich et al. and Doty et al. reported a decrease in extracellular matrix production in culture experiments with bone and cartilage cells and tumor cells under MG.^27,28^ In a parabolic flight experiment, the cytoskeletal systems such as actin and cytokeratin exposed to an MG environment were found to be altered and cells were observed to leave their adherent state, resulting in gene expression changes.^27,29^ Cytoskeletal changes, such as cytoskeleton loosening and microtubule shortening, in cells exposed to the MG environment have been reported in experiments in research space capsules and on the ISS.^9,30^ Moreover, we have previously shown that gravity influences several cell processes, including cell proliferation and differentiation, using MSCs cultured under MG environment.^1,11,12,25^ The MG environment inhibits the differentiation of hMSCs, rat and mouse MSCs, and human hematopoietic progenitor cells and promotes cell proliferation and survival.^8,11,25^ Additionally, an MG environment promotes the expression of neuroprotective and anti-inflammatory genes in cMSCs.^1^ These previous reports are consistent with the results of the in vitro experiments of the present study, explaining the upregulated features such as cell proliferation and angiogenesis and downregulated features such as microtubule and cell adhesion in the miRNA-MG group. Thus, in summary, the findings of this study provide novel evidence that an MG environment causes adaptive reorganization of the cytoskeleton in hcMSCs and changes cellular characteristics, thereby affecting nervous system-related genes and exerting multifaceted effects on cerebral infarction model, such as increased cell proliferation, angiogenesis, and synaptic transmission, resulting in better recovery of neurological function. Further studies, including clinical studies using hcMSCs cultured in the MG environment, and the accumulation of evidence are necessary. In future, hcMSCs established from free bone fragments obtained during craniotomy may be cultured in an MG environment for clinical application in patients with severe stroke requiring decompression craniotomy. Furthermore, determining efficient methods for culturing and establishing hcMSCs in an MG environment will enable the clinical application of hcMSCs in patients other than those with severe cerebral infarction.

This study had several limitations. First, the sample size was limited, especially for the gene-protein analysis, possibly because cerebral infarction in the rat model evaluated herein was relatively less severe, which minimized mortality. Second, although two evaluators who were blinded to cell administration groups independently assessed neurological function to minimize bias, there was a potential for bias.

In conclusion, hcMSCs cultured in a simulated MG environment may be a useful source of stem cells for restoring neurological function after cerebral infarction.

## Acknowledgments

We thank Takashi Otsuka and Takeshi Imura from the Division of Bio-Environmental Adaptation Sciences, Graduate School of Biomedical & Health Sciences, Hiroshima University for technical assistance. RNA sequencing, gene expression analysis, and GO analysis were supported by DNAFORM (Yokohama, Kanagawa).

## Data Availability

Data will be provided on request.

## Non-standard Abbreviations and Acronyms

1G: normal gravity
ANGPT1: angiopoietin 1
BDNF: brain-derived neurotrophic factor
bMSCs: bone marrow-derived mesenchymal stem cells
BSA: bovine serum albumin
cMSCs: cranial bone-derived mesenchymal stem cells
CNS: central nervous system
CTSS: cathepsin S
DCTN1: dynactin subunit 1
DWI: diffusion-weighted image
EPGN: epithelial mitogen
FBN2: fibrillin 2
FGF-7: fibroblast growth factor 7
Fgf2: fibroblast growth factor 2
Gap43: growth-associated protein 43
GDNF: glial cell-derived neurotrophic factor
GO: gene ontology
hbMSCs: human bone marrow-derived mesenchymal stem cells
hcMSCs: human cranial bone-derived mesenchymal stem cells
HE: hematoxylin-eosin
IBZ: ischemic border zone
ISS: International Space Station
ITGB3: integrin subunit beta 3
LAMA1: laminin subunit alpha 1
MCAO: middle cerebral artery occlusion
MG: microgravity
miRNA: microRNA
mNSS: modified neurological severity score
MRI: magnetic resonance imaging
MSCs: mesenchymal stem cells
Ngf: nerve growth factor
Ntrk2: neurotrophic tyrosine kinase receptor type 2
PBS: phosphate-buffered saline
PCR: polymerase chain reaction
PFA: paraformaldehyde
Sort1: sortilin 1
Act-b: β-Actin
Syp: synaptophysin
T2WI: T2-weighted image
VEGF-A: vascular endothelial growth factor-A
VEGF: vascular endothelial growth factor

## REFERENCES

1. Otsuka T, Imura T, Nakagawa K, Shrestha L, Takahashi S, Kawahara Y, Sueda T, Kurisu K, Yuge L. Simulated microgravity culture enhances the neuroprotective effects of human cranial bone-derived mesenchymal stem cells in traumatic brain injury. Stem Cells Dev. 2018;27:1287–1297. doi:10.1089/scd.2017.0299

2. Oshita J, Okazaki T, Mitsuhara T, Imura T, Nakagawa K, Otsuka T, Kurose T, Tamura T, Abiko M, Takeda M, et al. Early transplantation of human cranial bone-derived mesenchymal stem cells enhances functional recovery in ischemic stroke model rats. Neurol Med Chir (Tokyo). 2020;60:83–93.doi:10.2176/nmc.oa.2019-0186

3. Abiko M, Mitsuhara T, Okazaki T, Imura T, Nakagawa K, Otsuka T, Oshita J, Takeda M, Kawahara Y, Yuge L, et al. Rat cranial bone-derived mesenchymal stem cell transplantation promotes functional recovery in ischemic stroke model rats. Stem Cells Dev. 2018;27:1053–1061. doi:10.1089/scd.2018.0022

4. Cook D, Genever P. Regulation of mesenchymal stem cell differentiation. Adv Exp Med Biol. 2013;786:213–29. doi:10.1007/978-94-007-6621-1_12

5. Melief SM, Zwaginga JJ, Fibbe WE, Roelofs H. Adipose tissue-derived multipotent stromal cells have a higher immunomodulatory capacity than their bone marrow-derived counterparts. Stem Cells Transl Med. 2013;2:455–463. doi:10.5966/sctm.2012-0184

6. De Bari C, Dell’Accio F, Tylzanowski P, Luyten FP. Multipotent mesenchymal stem cells from adult human synovial membrane. Arthritis Rheum. 2001;44:1928–1942. doi:10.1002/1529-0131(200108)44:8<1928::Aid-art331>3.0.Co;2-p

7. Shinagawa K, Mitsuhara T, Okazaki T, Takeda M, Yamaguchi S, Magaki T, Okura Y, Uwatoko H, Kawahara Y, Yuge L, et al. The characteristics of human cranial bone marrow mesenchymal stem cells. Neurosci Lett. 2015;606:161–166. doi:10.1016/j.neulet.2015.08.056

8. Plett PA, Abonour R, Frankovitz SM, Orschell CM. Impact of modeled microgravity on migration, differentiation, and cell cycle control of primitive human hematopoietic progenitor cells. Exp Hematol. 2004;32:773–781. doi:10.1016/j.exphem.2004.03.014

9. Imura T, Otsuka T, Kawahara Y, Yuge L. ‘Microgravity’ as a unique and useful stem cell culture environment for cell-based therapy. Regen Ther. 2019;12:2–5. doi:10.1016/j.reth.2019.03.001

10. Imura T, Nakagawa K, Kawahara Y, Yuge L. Stem cell culture in microgravity and its application in cell-based therapy. Stem Cells Dev. 2018;27:1298–1302. doi:10.1089/scd.2017.0298

11. Yuge L, Sasaki A, Kawahara Y, Wu SL, Matsumoto M, Manabe T, Kajiume T, Takeda M, Magaki T, Takahashi T, et al. Simulated microgravity maintains the undifferentiated state and enhances the neural repair potential of bone marrow stromal cells. Stem Cells Dev. 2011;20:893–900. doi:10.1089/scd.2010.0294

12. Mitsuhara T, Takeda M, Yamaguchi S, Manabe T, Matsumoto M, Kawahara Y, Yuge L, Kurisu K. Simulated microgravity facilitates cell migration and neuroprotection after bone marrow stromal cell transplantation in spinal cord injury. Stem Cell Res Ther. 2013;4:35. doi:10.1186/scrt184

13. Ukai R, Honmou O, Harada K, Houkin K, Hamada H, Kocsis JD. Mesenchymal stem cells derived from peripheral blood protects against ischemia. J Neurotrauma. 2007;24:508–520. doi:10.1089/neu.2006.0161

14. Chopp M, Li Y. Treatment of neural injury with marrow stromal cells. Lancet Neurol. 2002;1:92–100. doi:10.1016/s1474-4422(02)00040-6

15. Han X, Qiu L, Zhang Y, Kong Q, Wang H, Wang H, Li H, Duan C, Wang Y, Song Y, et al. Transplantation of Sertoli-islet cell aggregates formed by microgravity: prolonged survival in diabetic rats. Exp Biol Med (Maywood). 2009;234:595–603. doi:10.3181/0812-rm-359

16. Mori S, Hatori N, Kawaguchi N, Hamada Y, Shih TC, Wu CY, Lam KS, Matsuura N, Yamamoto H, Takada YK, et al. The integrin-binding defective FGF2 mutants potently suppress FGF2 signalling and angiogenesis. Biosci Rep. 2017;37:BSR20170173. doi:10.1042/bsr20170173

17. Ornitz DM, Xu J, Colvin JS, McEwen DG, MacArthur CA, Coulier F, Gao G, Goldfarb M. Receptor specificity of the fibroblast growth factor family. J Biol Chem. 1996;271:15292–15297. doi:10.1074/jbc.271.25.15292

18. Einarsdottir E, Carlsson A, Minde J, Toolanen G, Svensson O, Solders G, Holmgren G, Holmberg D, Holmberg M. A mutation in the nerve growth factor beta gene (NGFB) causes loss of pain perception. Hum Mol Genet. 2004;13:799–805. doi:10.1093/hmg/ddh096

19. Carvalho OP, Thornton GK, Hertecant J, Houlden H, Nicholas AK, Cox JJ, Rielly M, Al-Gazali L, Woods CG. A novel NGF mutation clarifies the molecular mechanism and extends the phenotypic spectrum of the HSAN5 neuropathy. J Med Genet. 2011;48:131–135. doi:10.1136/jmg.2010.081455

20. Wiedenmann B, Franke WW. Identification and localization of synaptophysin, an integral membrane glycoprotein of Mr 38,000 characteristic of presynaptic vesicles. Cell. 1985;41:1017–1028. doi:10.1016/s0092-8674(85)80082-9

21. Otsuka T, Maeda Y, Kurose T, Nakagawa K, Mitsuhara T, Kawahara Y, Yuge L. Comparisons of neurotrophic effects of mesenchymal stem cells derived from different tissues on chronic spinal cord injury rats. Stem Cells Dev. 2021;30:865–875. doi:10.1089/scd.2021.0070

22. Nykjaer A, Lee R, Teng KK, Jansen P, Madsen P, Nielsen MS, Jacobsen C, Kliemannel M, Schwarz E, Willnow TE, et al. Sortilin is essential for proNGF-induced neuronal cell death. Nature. 2004;427:843–848. doi:10.1038/nature02319

23. Mikheeva IB, Shtanchaev RS, Pen’kova NA, Pavlik LL. Structure of interneuronal contacts in the neuropil of the oculomotor nuclei in mouse brain under conditions of long-term microgravity. Bull Exp Biol Med. 2018;165:457–460. doi:10.1007/s10517-018-4193-8

24. Mikheeva I, Mikhailova G, Shtanchaev R, Arkhipov V, Pavlik L. Influence of a 30-day spaceflight on the structure of motoneurons of the trochlear nerve nucleus in mice. Brain Res. 2021;1758:147331. doi:10.1016/j.brainres.2021.147331

25. Yuge L, Kajiume T, Tahara H, Kawahara Y, Umeda C, Yoshimoto R, Wu SL, Yamaoka K, Asashima M, Kataoka K, et al. Microgravity potentiates stem cell proliferation while sustaining the capability of differentiation. Stem Cells Dev. 2006;15:921–929. doi:10.1089/scd.2006.15.921

26. Monticone M, Liu Y, Pujic N, Cancedda R. Activation of nervous system development genes in bone marrow derived mesenchymal stem cells following spaceflight exposure. J Cell Biochem. 2010;111:442–452. doi:10.1002/jcb.22765

27. Ulbrich C, Pietsch J, Grosse J, Wehland M, Schulz H, Saar K, Hübner N, Hauslage J, Hemmersbach R, Braun M, et al. Differential gene regulation under altered gravity conditions in follicular thyroid cancer cells: relationship between the extracellular matrix and the cytoskeleton. Cell Physiol Biochem. 2011;28:185–198. doi:10.1159/000331730

28. Doty SB, Stiner D, Telford WG. The effect of spaceflight on cartilage cell cycle and differentiation. J Gravit Physiol. 1999;6:P89–P90.

29. Corydon TJ, Kopp S, Wehland M, Braun M, Schütte A, Mayer T, Hülsing T, Oltmann H, Schmitz B, Hemmersbach R, Gasset G, et al. Alterations of the cytoskeleton in human cells in space proved by life-cell imaging. Sci Rep. 2016;6:20043. doi:10.1038/srep20043

30. Vassy J, Portet S, Beil M, Millot G, Fauvel-Lafève F, Gasset G, Schoevaert D. Weightlessness acts on human breast cancer cell line MCF-7. Adv Space Res. 2003;32:1595– 1603. doi:10.1016/s0273-1177(03)90400-5

